# The Relationship between Noise Pollution and Depression and Implications for Healthy Ageing: A Spatial Analysis using Routinely Collected Primary Care Data

**DOI:** 10.1101/2024.07.15.24310019

**Authors:** Dialechti Tsimpida, Anastasia Tsakiridi

## Abstract

Environmental noise is a significant public health concern, ranking among the top environmental risks to citizens’ health and quality of life. Despite various studies exploring the effects of atmospheric pollution on mental health, spatial investigations into the effects of noise pollution have been notably absent. This study addresses this gap by investigating the association between noise pollution (from road and rail networks) and depression for the first time in England and first explores localised patterns based on area deprivation. Depression prevalence, defined as the percentage of patients with a recorded depression diagnosis was calculated in small areas within Cheshire and Merseyside ICS using the Quality and Outcomes Framework Indicators dataset for 2019. Strategic noise mapping for rail and road noise (LDEN) was employed to quantify noise pollution, indicating a 24-hour annual average noise level with distinct weightings for evening and night periods. The English Index of Multiple Deprivation (IMD) was utilised to represent neighbourhood deprivation. Geographical Weighted Regression and Generalised Structural Equation Spatial Modelling (GSESM) were applied to estimate relationships between transportation noise, depression prevalence, and IMD at the Lower Super Output Area (LSOA) level. While transportation noise showed a low direct effect on depression levels in Cheshire and Merseyside ICS, it significantly mediated other factors linked to depression prevalence. Notably, GSESM revealed that health deprivation and disability was strongly associated (0.62) with depression through the indirect effect of environmental noise, particularly where transportation noise exceeds 55 dB on a 24-hour basis. Comprehending variations in noise exposure across different areas is paramount. This research not only provides valuable insights for informed decision-making but also lays the groundwork for implementing noise mitigation measures. These measures are aimed at addressing mental health inequalities, enhancing the quality of life for the exposed population and supporting a healthier ageing process in urban environments. The findings also carry crucial implications for public health, specifically in tailoring targeted interventions to mitigate noise-related health risks in areas where noise burdens exceed 55 dB, and residents may experience health deprivation and disability.

## 1. Introduction

The increased demand for aircraft, road and railway transportation as a result of urbanisation has also increased noise pollution. Researchers, policymakers and urban planners have devoted great attention to this matter as noise pollution is regarded as the second greater environmental stressor impacting human health and well-being, after air pollution (Vienneau et al., 2015).

According to the World Health Organization, noise has been recognised as the top environmental risk to health (World Health Organization, 2018). It is estimated that around a hundred million people in the European Union are influenced by traffic-related noise, according to the EU’s Environmental Noise Directive, and traffic-related noise alone in Western Europe is responsible for almost 1.6 million healthy years of life lost each year (Dreger et al., 2019). Also, there have been indications that at least 1 million deaths every year in Western Europe are a result of traffic-related noise (Theakston and Weltgesundheitsorganisation, 2011).

In October 2022, the Journal of Mental Health editorial -an international forum for the latest research in the mental health field-quoted that there have been many studies that discussed the effects of the environment on mental health and have occasionally noted the possible ill effects of atmospheric pollution. A surprising omission, however, has been any discussion of the effects of noise pollution on mental health (Guha, 2022). The exact quote was that ‘the extent to which the effects of noise on mental health are omitted from research is irritating’. There are many studies where noise pollution has simply not been taken into account. Furthermore, a recent systematic literature review and meta-analysis revealed that the current evidence regarding the link between traffic noise and depression is of a “very low” quality (Dzhambov and Lercher, 2019).

In the UK, research exploring the impact of noise pollution on mental health is also scarce, (Guha, 2022) and given the existing mental health burden, this area of research presents a promising avenue for further investigation and for promoting the benefits of hearing conservation as a way to protect the population’s mental health. Furthermore, a review of the evidence in the WHO European Region revealed that the burden of noise seems to be unequally distributed in societies, calling for research on the social distribution of environmental noise exposure on a small spatial scale (Dreger et al., 2019).

Therefore, the aim of this study was a) to explore the link between noise pollution (from road and rail network) and depression in Cheshire and Merseyside Integrated Care System (ICS), and b) investigate potential localised patterns according to area deprivation.

## 2. Material and Methods

### 2.1 Data Sources

To quantify noise pollution, we used the strategic noise mapping for rail and road noise (LDEN) (Department for Environment, Food & Rural Affairs, 2019). LDEN indicates a 24-hour annual average noise level with separate weightings for the evening and night periods and we calculated the 24-hour annual average noise levels in small areas in Cheshire and Merseyside ICS.

Our primary outcome measure was depression prevalence, defined as the percentage of patients with a diagnosis of depression in their medical records. We calculated depression prevalence in small areas in Cheshire and Merseyside ICS in 2019 extracting respective values from the dataset on Quality and Outcomes Framework Indicators: Depression prevalence (QOF_4_12) Version 1.00 (Daras et al., 2023).

To represent deprivation within these smaller areas, we utilised the English Index of Multiple Deprivation (IMD). The IMD is a widely used statistic within the UK that provides measures of relative deprivation in small areas in England. In our analyses, we used the latest English Index of Multiple Deprivation (IMD 2019), which measures relative deprivation in small areas in England (McLennan et al., 2019).

All geospatial models employed in the study focused on the Lower Super Output Area (LSOA) as the unit of analysis. Cheshire and Merseyside (ICS) encompass 1,562 LSOAs, each with an average population of 1,500 individuals, according to data from the Office for National Statistics as of 21 March 2021. (Office for National Statistics, 2021) Furthermore, to facilitate comparisons among sub-Integrated Care Board locations in Cheshire and Merseyside ICS (Cheshire, Halton, Knowsley, Liverpool, South Sefton, Southport and Formby, St Helens, Warrington, Wirral), digital vector boundaries for Integrated Care Boards in England were incorporated in the analyses. (Office for National Statistics, 2023)

### 2.2 Analytical Approach

The percentage of road and rail noise coverage was calculated based on intensity in dB within each LSOA in Cheshire and Merseyside. We considered five categories based on the 24-hour annual average noise levels: 55-59.9 dB, 60-64.9 dB, 65-69.9 dB, 70-74.5 dB and ≥75 dB. We calculated noise separately for rail and road noise in each area. Additionally, to assess the overall impact of rail and road noise in each area, we combined the two noise databases and calculated the total road and rail noise coverage for the 24-hour annual average noise, considering noise levels exceeding 55 dB, and then subtracting the intersecting area.

The prevalence of depression was described with minimum and maximum values, central tendency measures (mean and median), and dispersion measures (range, and standard deviation).

We employed Geographical Weighted Regression and Generalised Structural Equation Spatial Modelling (GSESM) to estimate the relationship between transportation noise, depression prevalence and IMD. Mediation models (and the indirect effects) were tested with ordinary least squared (OLS) regression analyses using ArcGIS pro 2.9.2 OLS tool. We applied the four-step approach (Baron and Kenny, 1986) to examine several OLS regression analysis models and examined the significance of the coefficients at each step to find the best-fitting path models. Next, we calculate the significance of mediation in the structural equation modelling by computing the difference between two regression coefficients (Judd and Kenny, 1981). Exponentiated coefficients and summary statistics are presented for each stage.

Statistical significance was set at the 99% confidence level. Analyses were performed in ArcGIS Pro Version 2.9.2 (Esri Inc., 2021) using the following tools, in order of execution: Union, Tabulate Intersection, Spatial Join tool, the Analysis Toolbox and the Spatial Statistic Toolbox.

## 3. Results

**Table 1** shows the summary statistics for the percentage of road and rail noise coverage based on intensity in dB within each of the local authorities in Cheshire and Merseyside. We found that Knowsley had the highest coverage of areas exposed to road noise on a 24-hour basis, followed by Warrington. Halton had the highest coverage of areas exposed to rail noise on a 24-hour basis, followed by Warrington and Cheshire. Combining road and rail noise, we found that Warrington and Knowsley had the highest percentage of noise coverage, with over half of their respective areas exposed to transportation noise exceeding 55 dB on a 24-hour basis. **Figure 1** illustrates the correlation between areas with transportation noise levels of 55 dB or higher and the prevalence of depression, with brown denoting areas that experienced both high transportation noise and high depression prevalence in 2019.

**Figure 1.**
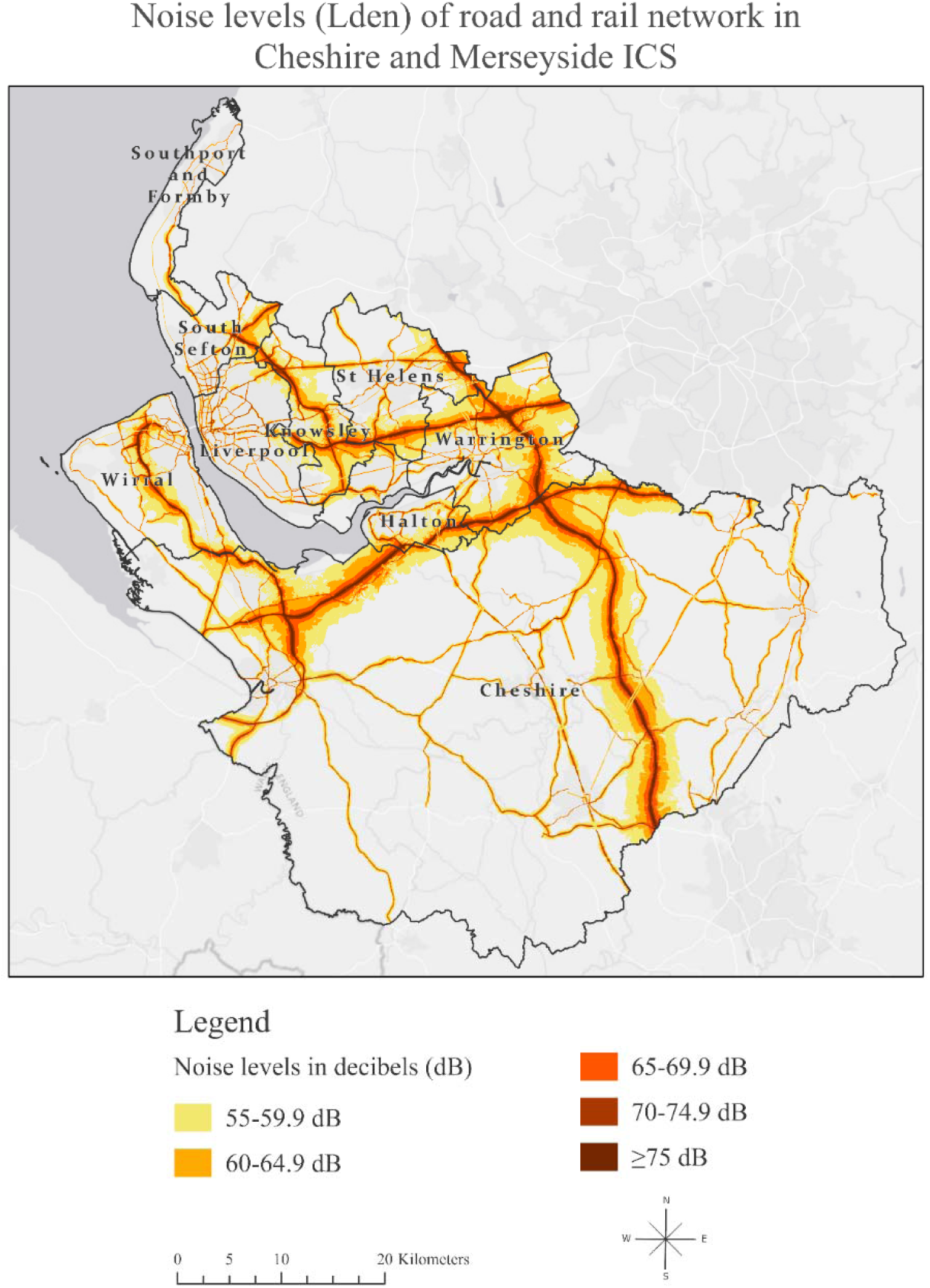
Noise levels of road and rail network in Cheshire and Merseyside ICS based on data derived from strategic noise mapping. (Department for Environment, Food & Rural Affairs, 2019)

**Table 1.** Summary statistics for the mean road and rail noise coverage percentage (%) and dB levels in Cheshire and Merseyside ICS in 2019.

| Sub ICB | 55-59.9 dB | 60-64.9dB | 65-69.9 dB | 70-74.9 dB | ≥75 dB | Total percentage of road and rail noise coverage with noise levels >55 dB subtracting their intersection |
| --- | --- | --- | --- | --- | --- | --- |
| Cheshire (mean road noise) | 11.57 | 5.60 | 2.95 | 1.85 | 0.67 | 24.69 |
| (mean rail noise) | 1.20 | 0.65 | 0.38 | 0.19 | 0.07 |  |
| Halton (mean road noise) | 17.13 | 8.24 | 4.37 | 2.68 | 1.56 | 36.69 |
| (mean rail noise) | 2.35 | 1.58 | 1.35 | 0.08 | 0.00 |  |
| Knowsley mean road noise) | 32.84 | 10.26 | 6.45 | 3.93 | 2.43 | 56.99 |
| (mean rail noise) | 1.74 | 1.12 | 0.85 | 0.01 | 0.00 |  |
| Liverpool mean road noise) | 11.17 | 4.75 | 3.26 | 3.25 | 1.01 | 24.96 |
| (mean rail noise) | 0.83 | 0.47 | 0.56 | 0.02 | 0.00 |  |
| South Sefton mean road noise) | 12.64 | 5.56 | 3.92 | 3.67 | 1.03 | 27.85 |
| (mean rail noise) | 0.87 | 0.53 | 0.00 | 0.00 | 0.00 |  |
| Southport and Formby (mean road noise) | 3.27 | 2.21 | 2.08 | 1.35 | 0.11 | 11.06 |
| (mean rail noise) | 1.15 | 0.99 | 0.00 | 0.00 | 0.00 |  |
| St Helens (mean road noise) | 17.04 | 6.05 | 3.51 | 2.44 | 0.95 | 31.24 |
| (mean rail noise) | 1.06 | 1.01 | 0.44 | 0.05 | 0.00 |  |
| Warrington (mean road noise) | 30.93 | 13.08 | 5.75 | 3.50 | 2.06 | 57.25 |
| (mean rail noise) | 1.65 | 1.03 | 0.80 | 0.15 | 0.00 |  |
| Wirral (mean road noise) | 13.87 | 6.05 | 3.90 | 2.96 | 1.28 | 28.72 |
| (mean rail noise) | 0.69 | 0.30 | 0.00 | 0.00 | 0.00 |  |

The summary statistics of depression prevalence in 2019 are shown in **Table 2**. Knowsley had the highest percentage of depression. Additionally, as we saw earlier, Knowsley also had the highest coverage of areas exposed to road noise on a 24-hour basis, along with the highest percentage of total noise coverage. In fact, over half of their respective area was exposed to transportation noise exceeding 55 dB on a 24-hour basis.

**Table 2.** Summary statistics of recorded depression prevalence in Cheshire and Merseyside ICS in 2019.

| Sub ICB | Number of LSOA | Mean | Median | Minimum | Maximum | Range | Standard Deviation |
| --- | --- | --- | --- | --- | --- | --- | --- |
| Cheshire | 446 | 11.62 | 11.54 | 5.03 | 16.09 | 11.06 | 2.05 |
| Halton | 79 | 14.66 | 15.48 | 10.42 | 19.01 | 8.59 | 2.27 |
| Knowsley | 98 | 15.92 | 16.36 | 9.63 | 20.74 | 11.10 | 2.66 |
| Liverpool | 298 | 12.74 | 12.47 | 7.95 | 21.48 | 13.53 | 2.38 |
| South Sefton | 111 | 12.96 | 13.03 | 8.33 | 17.99 | 9.66 | 2.27 |
| Southport and Formby | 78 | 11.26 | 12.12 | 8.44 | 16.10 | 7.66 | 2.32 |
| St Helens | 119 | 15.14 | 15.32 | 10.17 | 18.50 | 8.33 | 1.47 |
| Warrington | 127 | 12.55 | 12.65 | 7.11 | 18.20 | 11.10 | 3.00 |
| Wirral | 206 | 16.42 | 16.60 | 10.88 | 25.51 | 14.62 | 3.17 |

**Figure 2** illustrates the correlation between areas with transportation noise levels of 55 dB or higher and the prevalence of depression. Darker shades of purple represent higher depression prevalence, while darker shades of green indicate a greater extent of road and rail network noise coverage exceeding 55 dB in that region. We can also observe their combined interaction, with the brown colour denoting areas that experienced both high transportation noise and high depression prevalence.

**Figure 2.**
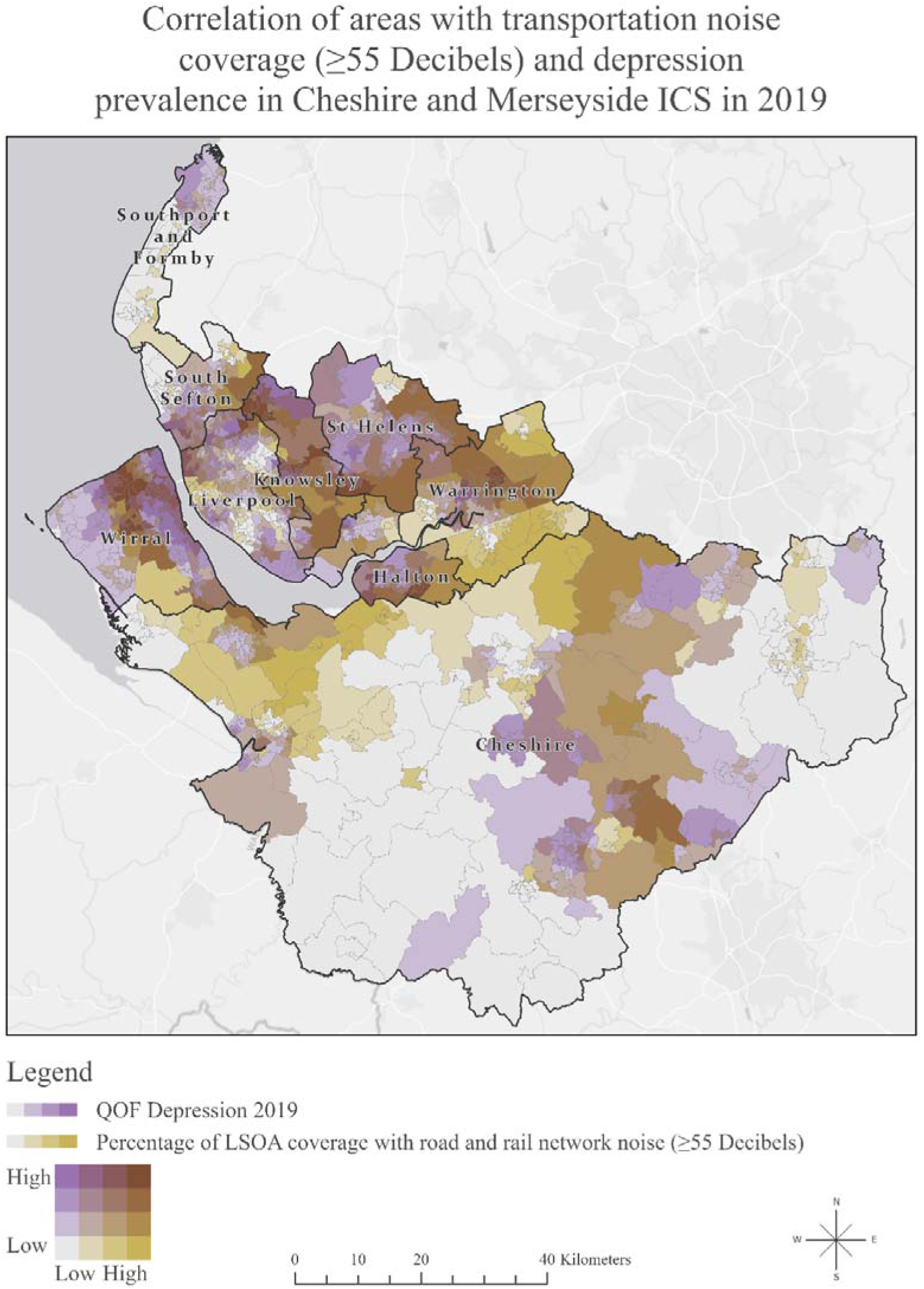
Map of Cheshire and Merseyside Integrated Care System showing the correlation of areas with transportation noise coverage (>55 Db) and depression prevalence in 2019.

**Figure 3** depicts the data on noise coverage 55 dB and above and depression in 2019 using the coefficients that show the correlation strength of the variables over space. The darker areas do not indicate where there is the highest noise pollution or highest depression prevalence; rather, they reveal where the relationship between noise pollution and depression is the strongest, informed by the results of Geographical weighted regression (GWR) (Naqvi et al., 2021). As we see, the local R-squared varies across the ICS, which shows that transportation noise may be a strong predictor of depression in one area, explaining up to 79% of the variance in depression in 2019 across the ICS. Summarised results of GWR between noise coverage and the prevalence of depression in 2019 in ICS are shown in **Table 3**.

**Figure 3.**
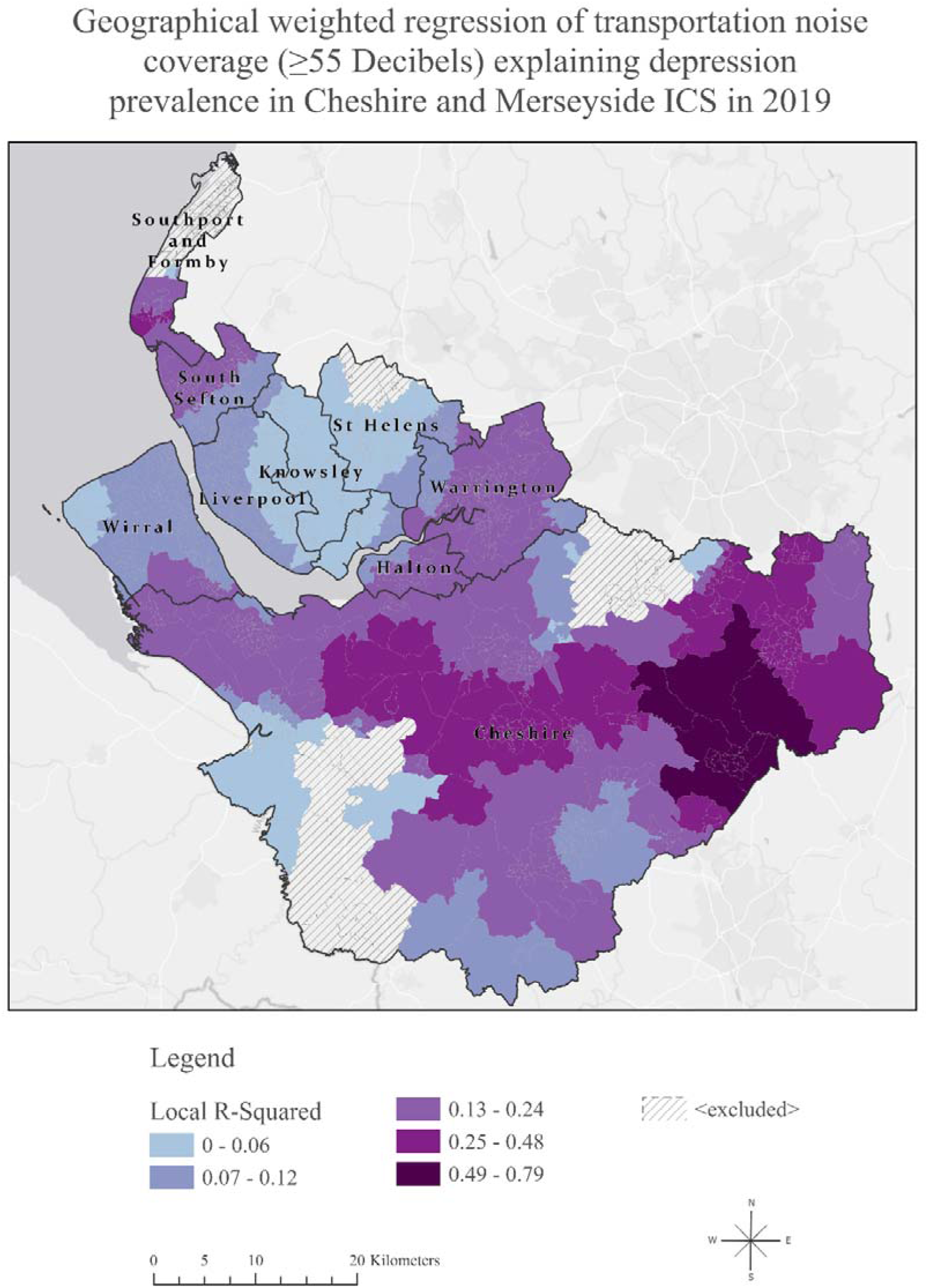
Geographically Weighted Regression Analysis: Examining the Relationship between transportation noise coverage and depression prevalence in Cheshire and Merseyside ICS, 2019.

**Table 3.**
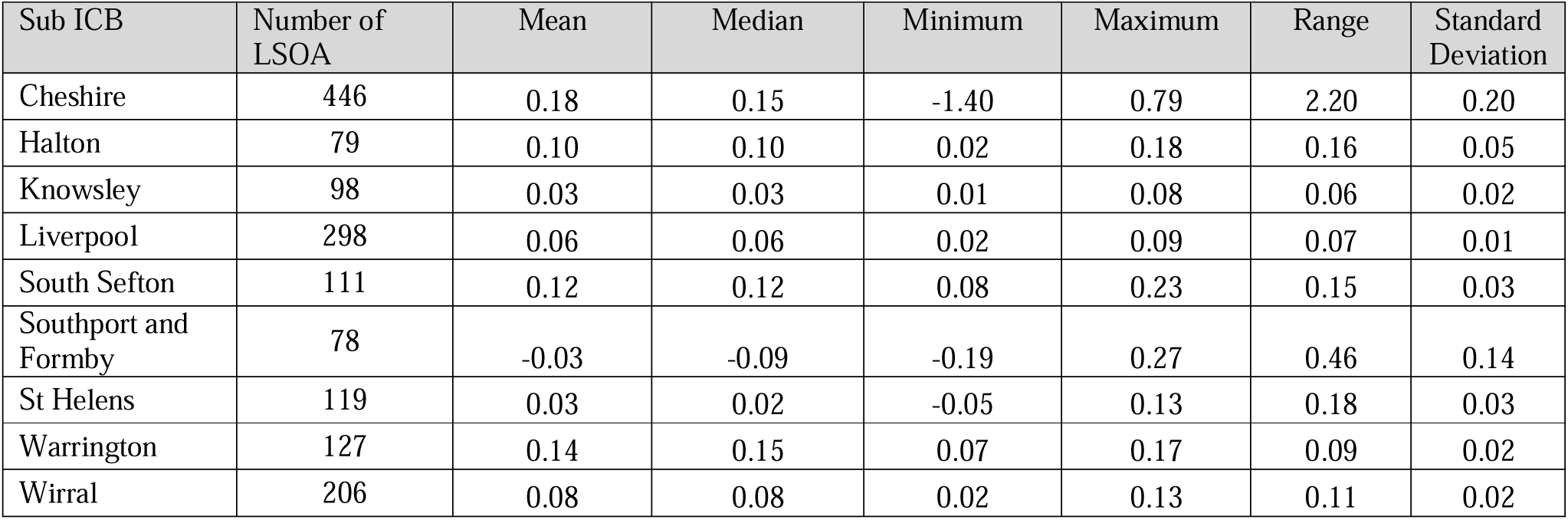
Summary statistics of the Geographically Weighted Regression Analysis: Exploring the Relationship between transportation noise coverage and depression prevalence in Cheshire and Merseyside ICS, 2019.

| Sub ICB | Number of LSOA | Mean | Median | Minimum | Maximum | Range | Standard Deviation |
| --- | --- | --- | --- | --- | --- | --- | --- |
| Cheshire | 446 | 0.18 | 0.15 | -1.40 | 0.79 | 2.20 | 0.20 |
| Halton | 79 | 0.10 | 0.10 | 0.02 | 0.18 | 0.16 | 0.05 |
| Knowsley | 98 | 0.03 | 0.03 | 0.01 | 0.08 | 0.06 | 0.02 |
| Liverpool | 298 | 0.06 | 0.06 | 0.02 | 0.09 | 0.07 | 0.01 |
| South Sefton | 111 | 0.12 | 0.12 | 0.08 | 0.23 | 0.15 | 0.03 |
| Southport and Formby | 78 | -0.03 | -0.09 | -0.19 | 0.27 | 0.46 | 0.14 |
| St Helens | 119 | 0.03 | 0.02 | -0.05 | 0.13 | 0.18 | 0.03 |
| Warrington | 127 | 0.14 | 0.15 | 0.07 | 0.17 | 0.09 | 0.02 |
| Wirral | 206 | 0.08 | 0.08 | 0.02 | 0.13 | 0.11 | 0.02 |

**Table 4** presents the results of GSESM. The analyses included the following four steps:

a) OLS regression analysis with all IMD domain scores predicting depression (step 1).
b) OLS regression analysis with all previously shown significant IMD domain scores predicting the percentage of environmental noise covered area per LSOA (step 2).
c) OLS regression analysis with the percentage of environmental noise covered area per LSOA predicting depression (step 3).
d) OLS significant IMD domain scores and environmental noise covered area per LSOA predicting depression (step 4)

**Table 4.** Standardised effects of the generalised structural equation spatial modelling (GSESM) mediation analyses of transportation in QOF-Depression 2019 in Merseyside.

| **Step 1** |  |  |  |  |
| --- | --- | --- | --- | --- |
| **IMD Domain Scores Predicting QOF-Depression** |  |  |  |  |
| **Variable** | **Coefficient<sup>a</sup>** | **StdError** | **Robust Pr<sup>b</sup>** |  |
| Income Deprivation | 11.673728 | 4.529014 | 0.005266* |  |
| Employment Deprivation | -3.623193 | 4.176416 | 0.385884 |  |
| Education Skills and Training | 0.024592 | 0.007433 | 0.000852* |  |
| Health Deprivation and Disability | 2.427641 | 0.201203 | 0.000000* |  |
| Crime | -0.032888 | 0.123761 | 0.7879 |  |
| Barriers to Housing and Services | -0.000566 | 0.007941 | 0.93813 |  |
| Living Environment Deprivation | -0.008936 | 0.004558 | 0.039140* |  |
| Income Deprivation Affecting Children Index | -2.215866 | 1.525089 | 0.091695 |  |
| Income Deprivation Affecting Older People Index | -12.597069 | 1.339679 | 0.000000* |  |
| **IMD Predicting Environmental Noise** |  |  |  |  |
| **Variable** | **Coefficient<sup>a</sup>** | **StdError** | **Robust Pr<sup>b</sup>** |  |
| Income Deprivation Domain | -84.985235 | 23.092033 | 0.000079* |  |
| Education Skills and Training | 0.296406 | 0.088988 | 0.000615* |  |
| Health Deprivation and Disability | 5.524358 | 2.041461 | 0.005514* |  |
| Living Environment Deprivation | -0.175043 | 0.051252 | 0.000082* |  |
| Income Deprivation Affecting Older People Index | 19.430965 | 14.423783 | 0.108445 |  |
| **Step 3** |  |  |  |  |
| **Environmental Noise per LSOA Predicting Depression** |  |  |  |  |
| **Variable** | **Coefficient<sup>a</sup>** | **StdError** | **Robust Pr<sup>b</sup>** |  |
| Environmental noise per LSOA | 0.015926 | 0.002496 | 0.000000* |  |
| **Step 4** |  |  |  |  |
| **IMD Domain Scores and Environmental noise per LSOA Predicting Depression** |  |  |  |  |
| **Variable** | **Coefficient<sup>a</sup>** | **StdError** | **Robust Pr<sup>b</sup>** |  |
| Income Deprivation | -2.327198 | 1.633573 | 0.24096 |  |
| Education Skills and Training | 0.021382 | 0.007307 | 0.005960* |  |
| Health Deprivation and Disability | 1.809736 | 0.161545 | 0.000000* |  |
| Living Environment Deprivation | -0.011395 | 0.004196 | 0.005322* |  |
| Environmental noise per LSOA | 0.011346 | 0.002074 | 0.000000* |  |
| **Calculation of Indirect Effect (Judd & Kenny Difference of Coefficients Approach)** |  |  |  |  |
| **Variable** | **Step 1 Coefficient** | **Step 4 Coefficient** | **Indirect effect of transportation on noise (step1-step4 coefficients)** | **Robust Pr<sup>b</sup>** |
| Education Skills and Training | 0.024592 | 0.021382 | 0.00321 | 0.000000* |
| Health Deprivation and Disability | 2.427641 | 1.809736 | 0.617905 | 0.000000* |
| Living Environment Deprivation | -0.008936 | -0.011395 | 0.002459 | 0.000000* |
| *An asterisk next to a number indicates a statistically significant p-value (p < 0.01).<br>a Coefficient: Represents the strength and type of relationship between each explanatory variable and the dependent variable.<br>b Robust Probability (Robust Pr): Asterisk (*) indicates a coefficient is statistically significant (p<0.01) |  |  |  |  |

Through Steps 1-4 we established that zero-order relationships among the variables exist, to establish that IMD and percentage of environmental noise covered area per LSOA have mediating effects.

The calculation of indirect effect through the Judd & Kenny Difference of Coefficients Approach (Judd and Kenny, 1981) suggested that although transportation noise had low direct effect in explaining depression levels in Cheshire and Merseyside ICS, it did significantly mediate other factors associated with depression prevalence. One of the most significant findings from the GSESM is the importance of noise in the effects of health deprivation and disability. Health deprivation and disability was strongly associated (0.62) with depression through the indirect effect of environmental noise where exceeding 55 dB LDEN.

## 4. Discussion

### 4.1 Summary of main findings

To the best of our knowledge, this is the first study to investigate the impact of transportation noise pollution on mental health in England. Our study identified areas with a heavier noise burden, offering the opportunity to tailor public health interventions in these regions to enhance the quality of life in urban environments. Combining transportation noise from road and rail networks, we found that Warrington and Knowsley had the highest percentage of noise coverage, with over half of their respective areas exposed to transportation noise exceeding 55 dB on a 24-hour basis.

Our research revealed that, although transportation noise did not have an equal direct role in explaining depression levels in all areas, it did play a significant mediating role, amplifying the effect of other factors on depression, such as the impact of health deprivation and disability.

### 4.2 Comparison with previous literature

Environmental noise pollution has been associated, in general with poor mental health indirectly (Dzhambov et al., 2017; Hammersen et al., 2016; Li et al., 2022) and the effects are varying from increased stress and anxiety levels (Gong et al., 2021; Jensen et al., 2018) to hyperactivity (Dreger et al., 2015; Haines et al., 2001), a decline of well-being (Schreckenberg et al., 2017) and an increase of psychotropic medication (Klompmaker et al., 2019; Okokon et al., 2018).

Previous studies investigating explicitly the association between environmental noise pollution and depression have yielded conflicting research results and an incomplete overview of the effects of all noise sources on mental health. As suggested by the systematic review and meta-analysis by Hegewald et al., which included 11 studies of road and 3 studies of railway traffic noise, there were indications of 2-3% increases in depression risk per 10 Db LDEN (Hegewald et al., 2020). Interestingly, a recent big data analysis using data from the UK Biobank found a negative association between moderate road traffic noise and major depression (Hao et al., 2022).

However, no previous study has examined local regression models to comprehend the varying relationships and spatial patterns, aiming to understand how local geographical factors influence the connections between variables, recognising that different places possess unique characteristics that impact these relationships. Moreover, although a US-based nationally representative survey explored the association between noise pollution and mental health in adolescents (Rudolph et al., 2019), no previous study has spatially examined the combined impact of road and rail noise on adults’ mental health, evaluating primary care records of depression as an outcome assessment.

### 4.3 Strengths and limitations

Our paper has a significant strength by employing an innovative structural equation spatial modelling methodology in small geographical areas. This approach allows for the examination of multiple mediators and links in the spatial chain during the model testing process. Additionally, the GSESM analysis furnishes crucial information on model fit, gauging the consistency of the hypothesised mediational model with the data and establishing zero-order relationships among variables. Notably, this methodology, which has not been previously utilised in the literature, leverages recent advancements in computing power, opening up new possibilities for the analysis and modelling of spatial data (Lin and Wen, 2022; Naqvi et al., 2021).

Our study, therefore, breaks new ground by exploring the link between environmental noise and the spatial epidemiology of depression using spatial analysis methods. This unique aspect forms a significant strength, particularly as no prior research has delved into this connection using spatial methodologies. This research contributes to the field by expanding the scope beyond earlier investigations limited to selected primary care practices (Al-Amoud et al., 2023). Instead, our study encompasses the entirety of practices in the Cheshire and Merseyside Integrated Care System (ICS), analysing records from a substantial 2.7 million individuals. The application of spatial methodology empowers us to identify spatial clustering patterns, pinpoint localized hotspots, and discern specific local risk factors—accomplishments that traditional non-spatial regression models are not achieving.

However, it is crucial to acknowledge several limitations. The data on depression are reliant on the recording practices of General Practitioners (GPs), introducing a potential source of bias. GPs’ decisions to diagnose depression may also be influenced by personal biases or preferences, as they might record symptoms instead of providing a formal diagnosis of depression (Grigoroglou et al., 2020). Another constraint is that the Quality and Outcomes Framework (QOF) depression prevalence offers aggregate data for all adults without specific information on adolescents or distinct age groups, nor details on those in retirement status, potentially confounding the associations (Tsimpida, Kontopantelis, et al., 2022).

### 4.4 Research and policy implications

The findings from this research carry significant implications for public health and the promotion of a healthy ageing process, which involves sustaining functional ability to ensure well-being.

Understanding the extent of noise exposure in various local authorities, particularly in areas where transportation noise exceeds 55 dB on a 24-hour basis, is essential. This study goes one step further to pinpoint areas with a heavier noise burden, allowing for the tailoring of public health interventions in these regions to enhance the quality of life for older residents and support a healthier ageing process.

Overall, this research offers a valuable foundation for informed decision-making and targeted strategies to reduce noise-related health risks in affected local authorities, ultimately contributing to the well-being and healthy ageing of the population.

Future research should explore the spatial relationship between noise pollution and other health outcomes, including cardiovascular disease (Münzel et al., 2016; Vienneau et al., 2015), sleep disturbance (Rudolph et al., 2019; Sygna et al., 2014; Van Kempen and Babisch, 2012; Zaharna and Guilleminault, 2010), cognitive decline (Haines et al., 2001; Stansfeld & Clark, 2015; Tzivian et al., 2016), metabolic syndrome (Yu et al., 2020), diabetes (Zare Sakhvidi et al., 2018), obesity (Wang et al., 2021), dementia (Cantuaria et al., 2021) and tinnitus (Theakston and Weltgesundheitsorganisation, 2011)

## Conclusion

While numerous studies explore the impact of the environment on mental health and occasionally acknowledge the potential adverse effects of atmospheric pollution, a notable omission has been the absence of discussions regarding the effects of noise pollution on mental well-being. Our study addresses this gap by providing novel insights into the correlation between noise pollution and mental health and, first, revealing the impact of noise pollution as a mediator, exaggerating the impact of health deprivation and disability on depression.

This research establishes a crucial foundation for informed decision-making and the development of targeted strategies to mitigate noise-related mental health risks in affected local authorities where high risk group reside. Ultimately, this contribution aims to tackle mental health inequalities, address their widening and promote healthy ageing in the population.

## Author statement

DT and AT made substantial contributions to the design of the work and were responsible for conducting the analyses, mapping, and drafting the manuscript. DT and AT contributed equally to this work and should be regarded as joint first authors. Both authors substantially contributed to the interpretation of data for the work and revised it critically for important intellectual content. Both authors have read and approved the final manuscript and agree to be accountable for all aspects of the work in ensuring that questions related to the accuracy or integrity of any part of the work are appropriately investigated and resolved.

## Data Availability

The dataset on Quality and Outcomes Framework Indicators: Depression prevalence (QOF_4_12) is publicly available in the Place-based longitudinal data resource (Original record link: https://pldr.org/dataset/2ldz5, Data catalogue DOI: 10.17638/datacat.liverpool.ac.uk/2170).

## Acknowledgements

This research was funded by the Institute of Population Health at the University of Liverpool and was supported by the Centre for Research on Ageing at the University of Southampton and the UK Hearing Conservation Association.

